# A Comprehensive Approach to Days’ Supply Estimation in a Real-World Prescription Database: Data Cleaning, Imputation, and Adherence Analysis

**DOI:** 10.1101/2025.09.03.25335007

**Authors:** Maria Malk, Kerli Mooses, Marek Oja, Johannes Holm, Hanna Keidong, Nikita Umov, Sirli Tamm, Sulev Reisberg, Jaak Vilo, Raivo Kolde

**Affiliations:** Institute of Computer Science, University of Tartu, Estonia; The Institute of Family Medicine and Public Health, University of Tartu, Estonia

**Keywords:** prescription records, days’ supply, medication adherence, CMA, daily dose, imputation

## Abstract

**Background:** For accurate medication usage statistics and medication adherence calculations, we need to have an accurate days’ supply (DS) for each prescription. Unfortunately, often the DS or information needed for calculating the DS is not provided. Therefore, other methods need to be applied to acquire missing values or substituting incorrect values.

**Objective:** The aim of this study is to apply a variety of methods for managing incomplete and missing data to enhance the accuracy of calculating DS for all medications and drug forms alike. Furthermore, to describe the effect of applied methods on the medication adherence calculated on real-world data.

**Methods:** A dataset comprising prescription records from a 10% random sample of the Estonian population between 2012 and 2019 was used. The workflow consisted of three steps – data cleaning, imputation and calculation of DS. For imputation, different methods were combined, such as calculating mode-based daily dose, or using usage guidelines from Summary of Product Characteristics (SPCs) or legislation. DS was calculated based on provided daily dose or imputed value. To evaluate the impact of data cleaning, medication adherence for baseline dataset and corrected dataset for two time periods 2012–2015 and 2017–2019 was calculated and compared.

**Results:** The drug forms with the lowest proportion of correct DS provided were insulin injections (3.1%) and intravaginal contraceptives (8.0%) while the highest proportion of DS was provided for inhalation medication (57.5%), oral drops (53.0%) and tablets, capsules, suppositories (45.8%). As a result of applying different imputation approaches, we successfully found the DS for 98.3% (N=7,415,347) of dispensed prescriptions. For the remaining 1.7% (N=129,545) of prescriptions DS could not be imputed nor calculated with these methods. As for the medication adherence, the distinction between two observed time periods was more distinct in the baseline dataset compared with the corrected dataset for most of the drug groups, indicating that the applied correction methods had lessened the stark contrast.

**Conclusions:** In summary, our study demonstrated that with a carefully designed imputation pipeline where data-driven imputation is combined with domain knowledge and literature information, it is possible to meaningfully improve the quality of prescription datasets and generate more accurate and consistent adherence metrics across various drug form. Nonetheless, future efforts should continue to refine imputation techniques, incorporate machine learning approaches where appropriate, and expand validation efforts using external benchmarks or clinical outcomes.

## Introduction

Electronic healthcare databases provide a valuable data source for conducting various studies, as they are detailed, structured, and often cover a long time span. One important source of such data is pharmacy medication records, which provide the opportunity to research medication usage and adherence cost-effectively and at scale [1-3]. To calculate medication adherence, an accurate days’ supply (DS) for each prescription is needed [4]. DS describes for how many days the dispensed medication is expected to last. In medical fields, DS is also referred to as treatment course length. While some prescription databases have DS recorded, in others, the DS value needs to be calculated using other available information such as the number of dispensed medications and prescribed daily dose [4]. Unfortunately, there is an abundance of evidence suggesting inaccuracies and missing values in the prescription databases [4-5].

Several studies have specifically addressed the challenges related to missing daily dose and DS issue [4, 6-7]. For example, imputing one dose of medication per day for missing daily doses has been shown to work for stroke patients [6], while imputing mode daily dose per active substance and number of tablets per prescriptions has been shown to work for diabetic drugs [4]. Studies have also demonstrated that daily dose values can be imputed using machine learning algorithms that incorporate various patient characteristics [4]. Some studies [8-9] have applied defined daily dose (DDD) toolkit developed by World Health Organisation [10]. However, it has been concluded that using DDD as a daily dose substitute, may lead to misclassification of medication adherence [8-9]. The limitations of the existing studies tackling the missing daily dose and DS issue are that they often focus on one disease or medication group [4, 6-7, 9, 11-13] and thus, it is unknown whether the same approach is applicable to other active substances or diseases. Moreover, studies have often been conducted using single-dose oral medications [4, 6-7, 9] and little is known how to address the missing data in other drug forms such as eye drops, topical cremes and gels. Although some studies have researched medication adherence among diseases that often use other drug forms [11, 13-18], only a few of these utilise pharmacy records [13, 17]. However, these studies have not tackled the problem of missing data.

In addition to missing data, some studies have highlighted that some inaccuracies may be present in DS values [5, 19]. More common inconsistencies are in reported DS values, dosage, fill intervals, administration times and quantity [5, 19]. It has been stressed that further research is needed to evaluate DS reporting errors and to recommend strategies to address these errors [20].

To the best of our knowledge, no comprehensive approach exists that addresses both missing as well as inaccurate data in prescriptions database across all prescriptions, irrespective of the active substance or drug form. To address this shortcoming, the aim of this study is to apply a variety of methods for managing incomplete and missing data to enhance the accuracy of calculating DS for all medications and drug forms alike. In addition, we describe the effect of applied methods on the medication adherence calculated on real-world data.

## Methods

### Data

The dataset used in this study consisted of prescription data for a 10% random sample of the Estonian population, covering the period from 2012 to 2019 [21]. The data originated from national e-prescription database, where all prescriptions issued in primary and secondary care are stored since 2010. The dataset includes all prescribed medications together with their dispensing information. Specific information about the prescriptions is shown in Table 1. Notably, the dataset does not contain information about over-the-counter medications nor inpatient medications.

**Table 1.** Available prescription information.

| Category | Details |
| --- | --- |
| Patient information | Unique ID for each patient |
| General administrative information | Unique ID for each prescription |
|  | Issue date |
|  | Validity date |
|  | Dispensed date |
|  | Prescription type (initial, refill, narcotic, medical device) |
|  | Prescribing healthcare provider information |
|  | Diagnosis code for which the medication was prescribed |
|  | Rationale for brand-specific prescribing |
|  | Dispensing pharmacy information |
|  | Prescription cancellation reason |
| Active substance | Name(s) of active substance |
|  | Amount(s) of max three components |
|  | Unique ATC<sup>a</sup> code for active substance |
| Drug form | Type of medication (e.g., tablet, cream, eye drops) |
| Daily dose information | Number of times the medication is taken or applied<sup>b</sup> |
|  | Amount of medication per dose<sup>b</sup> |
|  | Optional free-text |
| Days' supply | Optional days' supply |
| Dispensed package information | Unique ID for each drug package |
|  | Number of packages |
|  | Number of units in each package |
|  | Item size (for non-single-dose medications, e.g., creams, eye drops) |

<sup>a</sup> Abbreviation: Anatomical Therapeutic Chemical – ATC
<sup>b</sup> These multiplied, make up the daily dose

In total, the dataset initially comprised of 9,279,082 prescriptions, of which 7,544,892 (81.3%) were dispensed by patients. The rest of the prescriptions were prescribed but never dispensed. As it stands, only the prescriptions that were dispensed, were included in this study and equipped with DS value.

There were several data quality issues in the database that had to be addressed. Providing daily dose information became mandatory for the doctors in mid-2016. As a result, a 67.4% of prescriptions issued prior to this date lack daily dose information compared with 0.4% of prescriptions issued in 2017–2019. Throughout this study period it was optional for the doctors to specify the treatment course length for each prescription. This treatment course length is equivalent to the DS and could be used as a substitute or for comparison of calculated DS. The number of prescriptions with provided DS increased in time – in 2012–2016 15.2% of prescriptions had this information compared with 36.1% in 2017–2019.

Even though almost all prescriptions after 2016 were provided with a daily dose, the information was inconsistent in terms of used units. The amount of medication could be given as a quantity (e.g. 1 tablet, 2 pills, etc.) or the amount of active substance (e.g. 10 mg, 20 mg, etc.). Neither of these writing methods were consistent among themselves either. For example, if a drug contained two active substances, like 5 mg + 10 mg, and the doctor decided to write the amount taken at once in active substance amount, they had 3 distinct ways to write that: 5 mg, 10 mg and 15 mg.

For non-single-dose medications such as drops, creams, and gels, daily dose was often noted only qualitatively (e.g., ‘once per day’), without quantitative detail, making days’ supply impossible to calculate.

### Workflow

The main workflow to acquire the DS value is presented in Figure 1.

**Figure 1.**
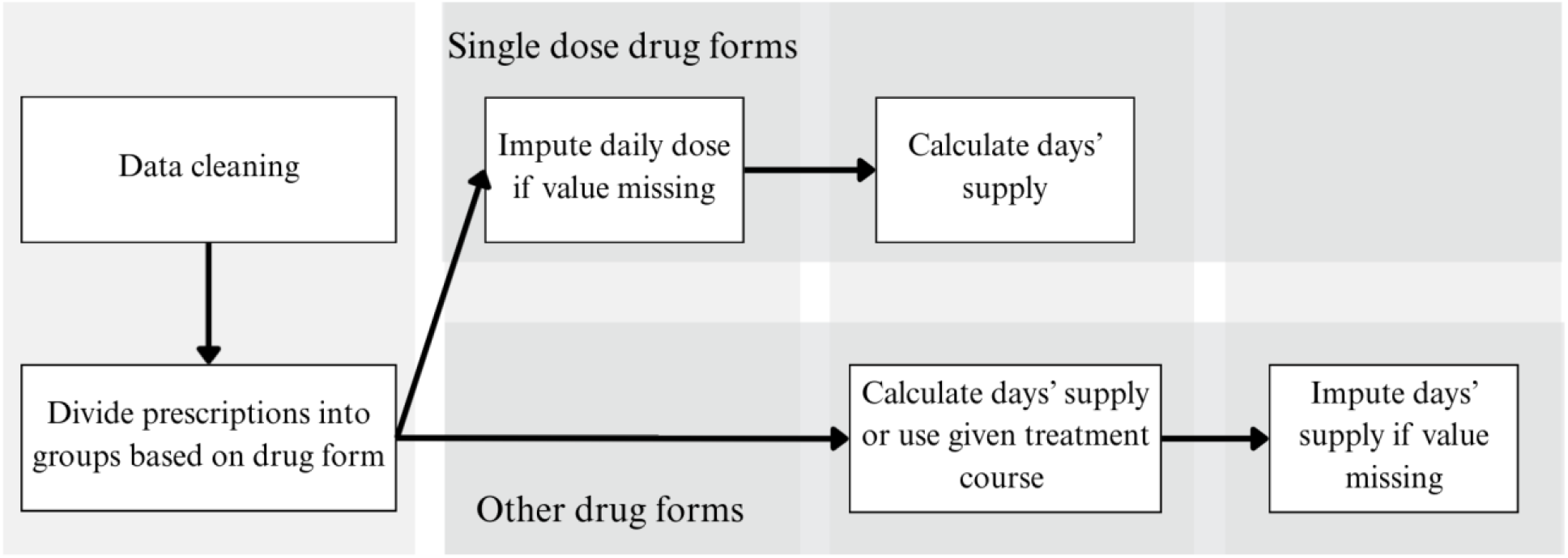
Workflow to acquire days’ supply.

#### Data Cleaning and Drug Form Grouping

The prescriptions were first checked for correctness. Some of the prescriptions from 2012 had false active substance data. Since every dispensed medication has a package number, the correct active substance was taken from package information.

Prescriptions were stratified by drug form into distinct groups, since each category follows specific conventions for stating the daily dose. For example, liquid formulations record daily doses in drops, creams in standardized dose units, and sprays in actuations. If needed, these groups were further divided by active substances because DS can be imputed more correctly for pharmacologically similar active substances and drug forms. For example, ophthalmic preparations were initially divided by drug form into two groups: single-dose containers and multiple-dose containers. The multiple-dose container group was further subdivided into two categories: one containing anti-infective, anti-inflammatory agents, and their combinations, and another containing all other substances. The last two subgroups required different imputation values.

All tablets, capsules, and suppositories share similar daily dose parameters, which allows for a consistent approach in calculating DS. These were collectively referred to as single-dose medications. Groups containing fewer than 1,000 prescriptions were given DS value based on the provided DS data and were excluded from further imputing processes.

#### Single Dose Drug Forms Days’ Supply

For single-dose medications, if all the necessary daily dose information was available, we calculated the DS with a method that took into consideration whether the daily dose was written in dose units (1 tablet, 2 tablets, etc) or active substance amount (5 mg, 10 mg, etc), since there are many different ways to prescribe the daily dose value. Afterwards, we compared the calculated value with provided DS, if it was available. Whichever of the two – calculated or provided DS – had smaller value, it was used. T The rationale is that a pharmacist may dispense a larger package (i.e., more tablets) if no package exactly matches the prescription. If the provided DS was not available, the calculated DS was used.

For prescriptions where DS could not be determined using the forementioned logic, a mode-based imputation method was used. A reference dataset was created with the most common daily dose for each active substance and corresponding dose strength. When two or more daily dose values were equally frequent, those active substances were excluded from imputation and their prescriptions remained unimputed. DS was then calculated for imputed prescriptions. This approach adds dose specificity to the active substance classification, making the DS value more reliable compared to direct DS imputation.

#### Other Drug Forms Days’ Supply

The general workflow for determining DS in the other dosage‐form groups was as follows. The target drug group was thoroughly analysed to identify trends in how daily doses are recorded and the specific characteristics of its treatment regimens. Where possible, the DS was calculated, and the existence of the prescribed DS was checked. If prescribed DS information was available, it was either used directly or compared to the computed DS value, and the more appropriate of the two was adopted.

In cases where neither of these approaches could be applied, alternative imputation strategies were used. These strategies were based on Summary of Product Characteristics (SPCs) [22] where the recommended DS is written and on relevant Estonian regulations [23], where for example the recommended prescription usage length is recorded. These outcomes were then reviewed by a domain expert and, if necessary, adjusted to reflect current practices in the field. Based on this evaluation, specific imputation rules were defined for each medication group and subsequently applied in the processing workflow.

More specifically:

- For semisolid medications consisting of cremes, gels, shampoos and ointments for topical usage, if the provided DS was available, that value was used. Otherwise, it was imputed as 30 days per package.
- Medicinal nail polish treatment should last for 3 to 12 months depending on the condition. When the provided DS was available, it was used. Otherwise, DS was calculated based of the package type. As only two package types were present in the dataset, the DS was either 210 days or 180 days.
- For eye drops provided DS was used, if available. Otherwise, due to shelf-life limits, DS was imputed as 30 days per package, except for anti-infectious, anti-inflammatory, and corticosteroid drops, which should not exceed a two-week treatment course. Therefore, DS were imputed with either 30 days or 14 days depending on the active substance. When the eye drops were packaged in a single dose container and daily dose was provided, daily dose was used to calculate DS. If daily does was not available, the provided DS was used and if neither was available, it was imputed as 30 days.
- For ear drops, all medication present in the dataset were for short time use only. Namely, anti-infective and analgetic medications, that are usually not used for more than 7 days without doctor supervision. When the provided DS was available, it was used. Otherwise, a 7-day value was used for imputation.
- For oral drops, if the daily dose was specified quantitatively (e.g., in number of drops or amount of active substance), this information was used to calculate the DS. The calculations varied depending on the active substance, as the drop sizes differed between substances. When the DS could not be calculated, and provided DS was available, it was used. Otherwise, 30 days per package was imputed due to shelf-life limits.
- For nasal sprays, DS was calculated based on the daily dose if it was provided. If the DS could not be calculated, we used provided DS if it was available. Otherwise, a default imputed value of 30 days per package was applied except for anti-fungal nasal spray, where the treatment course should not last more than 7 days. Therefore, for imputation, the 7-day value was used.
- For inhalation powders, if the medication was divided in blisters and daily dose was provided, then it was used to calculate DS. When the DS could not be calculated, and provided DS was available, it was used. Otherwise, a value of 60 days was imputed for all prescriptions, considering the prescription guidelines and prevailing trends. According to Estonian legislation and prescribing practices, chronic medications are issued on a single prescription for a two-month period.
- For syrups, if the daily dose was specified quantitatively, it was used to calculate the DS. When the DS could not be calculated, and provided DS was available, it was used. Otherwise, 30 days per package was imputed due to shelf-life limits.
- For antibiotic solutions, when the provided DS was available, it was used. For these medications, the treatment course should not be longer than 14 days, therefore, for imputation 14-day value was used.
- For transdermal patches, when the provided DS was available, it was used. Otherwise, the DS was calculated based on the length of the effect of the patches based on information from SPCs.
- For vaccines and implants, the treatment course was imputed as 1 day.
- For insulin, a value of 60 days was imputed for all prescriptions, considering the prescription guidelines and prevailing trends. According to Estonian legislation and prescribing practices, chronic medications are issued on a single prescription for a two-month period.

#### Quality Control Using Medication Adherence

To assess the impact of data cleaning and imputation, we calculated medication adherence for 147 active substances used in chronic conditions in both the baseline and corrected datasets. This approach provides a broader statistical perspective across all medications, allowing us to evaluate the validity of the imputations based on all the prescriptions available.

The active substances used in the analysis were selected as follows: 300 of the most frequently prescribed active substances were extracted from the database and two pharmacists independently filtered out those that were meant for chronic conditions. The active substances used in calculations were divided into 27 groups based on the ATC therapeutic subgroup (Appendix 1).

In the baseline dataset, the DS was calculated based on the provided daily dose for single-dose medications. When the DS was not provided, a value of 30 days was imputed [4]. Baseline dataset analyses were performed using a simplified approach, which is fast, inexpensive, and requires minimal data processing. In the corrected dataset, we applied more resource-intensive methods to evaluate whether such refinement provides a meaningful advantage in adherence estimation.

For medication adherence calculations, the continuous multiple interval measures of medication availability measure (CMA)[2] were used. Out of eight CMAs, CMA5 was selected as it accounts for gaps in medication availability and assumes that the new refill is stored until the previous prescription is fully used. The adherence was calculated on yearly basis. The calculation window began with the first medication dispensing and ended with the last, requiring each patient to have at least two prescriptions dispensed. Any unused medication remaining at the end of the window was excluded from the calculations [2]. The CMA implementation in AdhereR [24] was used through AdherenceFromOMOP [25].

The average CMA per year was calculated for both datasets – baseline and corrected. In addition, in both datasets two time periods were compared – 2012–2015 when providing daily dose information was voluntary and 2017–2019 when this information was mandatory for the doctors. Since 2016 was a transition year, it was excluded from the time-period comparison but is shown in the CMA graphs in Appendix 1. To describe the change in medication adherence between two periods, the change in period means was calculated.

The study was approved by the Research Ethics Committee of the University of Tartu (300/T-23) and the Estonian Committee on Bioethics and Human Research (1.1-12/653), and the requirement for informed consent was waived.

## Results

A total of 7,544,892 dispensed prescriptions were included into the process of establishing DS value. The largest drug form group was single-dose medications – including tablets, capsules, and suppositories – which accounted for 81.9% (N = 6,176,585) of all dispensed prescriptions. The remaining drug form groups are listed in Table 2. In total, 13 major drug form categories were identified.

**Table 2.** Prescription distribution by drug groups and days’ supply establishing methods.

| DRUG GROUP | DATASET INFORMATION | METHOD TO FINDING DAYS' SUPPLY | PRESCRIPTIONS IN MAIN GROUP |
| --- | --- | --- | --- |
| Tablets, capsules, suppositories | Daily dose provided | Calculated using given daily dose | 2 825 867 (45.75%) |
|  | Days' supply provided | Days' supply was used | 864 091 (13.99%) |
|  | Daily dose nor days' supply provided | Calculated using imputed daily dose value | 2 485 182 (40.23%) |
|  | Days' supply could not be calculated nor imputed |  | 1 445 (0.02%) |
| Semisolid dosage forms | Days' supply provided | Days' supply was used | 119 748 (37.55%) |
|  | Days' supply not provided | Imputed as 30 days per package<sup>a</sup> | 199 154 (62.45%) |
| Medicinal nail polish | Days' supply provided | Days' supply was used | 3 008 (41.55%) |
|  | Days' supply not provided | Imputed as 210 days or 180 days<sup>a</sup> | 4231 (58.45%) |
| Ear drops | Days' supply provided | Days' supply was used | 5 661 (57.11%) |
|  | Days' supply not provided | Imputed as 7 days per prescription<sup>a</sup> | 4 252 (42.89%) |
| Eye drops | Eye drops in single dose containers - daily dose provided | Calculated using given daily dose | 6 724 (2.35%) |
|  | Other eye drops - days' supply provided | Days' supply was used | 62 332 (21.81%) |
|  | Other eye drops - days' supply not provided | Imputed as 14<sup>a</sup> or 30<sup>b</sup> days per package | 216 740 (72.36%) |
|  | Days' supply not calculated nor imputed |  | 43 (0.02%) |
| Oral drops | Daily dose provided | Calculated using given daily dose | 52 085 (53.03%) |
|  | Days' supply provided | Days' supply was used | 33 055 (33.65%) |
|  | Daily dose nor days' supply provided | Imputed as 30 days per prescription<sup>a</sup> | 12 961 (13.20%) |
|  | Days' supply not calculated nor imputed |  | 120 (0.12%) |
| Inhalation medication | Daily dose provided | Calculated using given daily dose | 78 541 (57.50%) |
|  | Days' supply provided | Days' supply was used | 19 885 (14.56%) |
|  | Days' supply not provided | Imputed as 60 days per prescription<sup>a</sup> | 27 137 (27.19%) |
|  | Days' supply not calculated nor imputed |  | 1 025 (0.75%) |
| Nasal sprays | Anti-fungal nasal spray - days' supply not provided | Imputed as 7 days per prescription<sup>a</sup> | 7 738 (8.72%) |
|  | Daily dose provided | Calculated using given daily dose | 26 361 (29.70%) |
|  | Days' supply provided | Days' supply was used | 40 551 (45.69%) |
|  | Daily dose nor days' supply provided | Imputed as 30 days per prescription<sup>a</sup> | 14 094 (15.88%) |
| Syrups | Antibiotics - days' supply not provided | Imputed as 14 days per prescription<sup>a</sup> | 29 130 (34.31%) |
|  | Daily dose provided | Calculated using given daily dose | 37 251 (43.87%) |
|  | Days' supply provided | Days' supply was used | 3 094 (3.64%) |
|  | Days' supply not provided | Imputed as 30 days per package<sup>b</sup> | 15 436 (18.18%) |
| Transdermal patch | Hormonal patch - days' supply not provided | Impute as 7 days per patch plus 7 days<sup>a</sup> | 13 131 (81.18%) |
|  | Analgesic patch - days' supply not provided | Imputed as 3 days per patch<sup>a</sup> | 1 127 (7.01%) |
|  | Days' supply provided | Days' supply was used | 599 (5.94%) |
|  | Days' supply not provided | Impute as 7 days per patch<sup>a</sup> | 865 (5.38%) |
| Intravaginal contraceptives | Days' supply provided | Days' supply was used | 1 692 (8.00%) |
|  | Days' supply not provided | Imputed as 30 days per item in package<sup>a</sup> | 19 453 (92.00%) |
| Implants and vaccines | Days' supply provided | Days' supply was used | 18 940 (45.2%) |
|  | Days' supply not provided | Imputed as 1 day | 22 979 (54.8%) |
| Insulin injections | Days' supply provided | Days' supply was used | 2 601 (3.14%) |
|  | Days' supply not provided | Imputed as 60 days per prescription<sup>a</sup> | 80 266 (96.86%) |
| Other | Days' supply provided | Days' supply was used | 49 046 (27.88%) |
|  | Days' supply not calculated nor imputed |  | 126 895 (72.12%) |

<sup>a</sup> based on (SPC) issued by Estonian Agency of Medicines, <sup>b</sup> medication not shelf-stable for more than 30 days after opening

The drug forms with the lowest proportion of correct DS provided were insulin injections (3.1%) and intravaginal contraceptives (8.0%) while the highest proportion of DS was provided for inhalation medication (57.5%), oral drops (53.0%) and tablets, capsules, suppositories (45.8%) (table 2).

For tablets, capsules, and suppositories, 40.2% of prescriptions lacked daily dose information and were therefore imputed using the mode-based imputation described earlier. The mode table consisted of 1002 active substance and dosage amount combinations, out of which 60.7% of combinations had the value of once per day and 24.7% of these combinations had the value of twice per day. In cases where both a calculated DS and provided DS information were available, the smaller value was used out of the two. As a result, 14.0% of prescriptions in single-dose medications were assigned DS provided by the doctor. For other drug forms, when the DS could not be calculated, instead of using daily dose, we imputed the DS based on the specific drug form. The proportion of imputed prescriptions ranged from 13.2% for oral drops to 100% for implants, vaccines and intravaginal contraceptives (table 2). As a result of applying different imputation approaches, we successfully found the DS for 98.3% (N = 7,415,347) of dispensed prescriptions. For 1.7% (N = 129,545) of prescriptions DS could not be imputed nor calculated.

To evaluate the impact of data cleaning and imputation. the medication adherence of 147 active substances belonging to 27 ATC therapeutic subgroups were calculated. For most of the drug groups, the difference between two observed time periods was more distinct in the baseline dataset compared with the corrected dataset, indicating that the applied correction methods had lessened the stark contrast (Figure 2, Appendix 1). For the multiple dose medications, such as drugs for obstructive airway diseases (R03) and ophthalmologicals (S01), there was no distinction between the observed time periods. However, the medication adherence improved similarly for both time periods. At the same time, some adherence measures, such as the thyroid therapy (H03), showed 100% of medication adherence.

**Figure 2.**
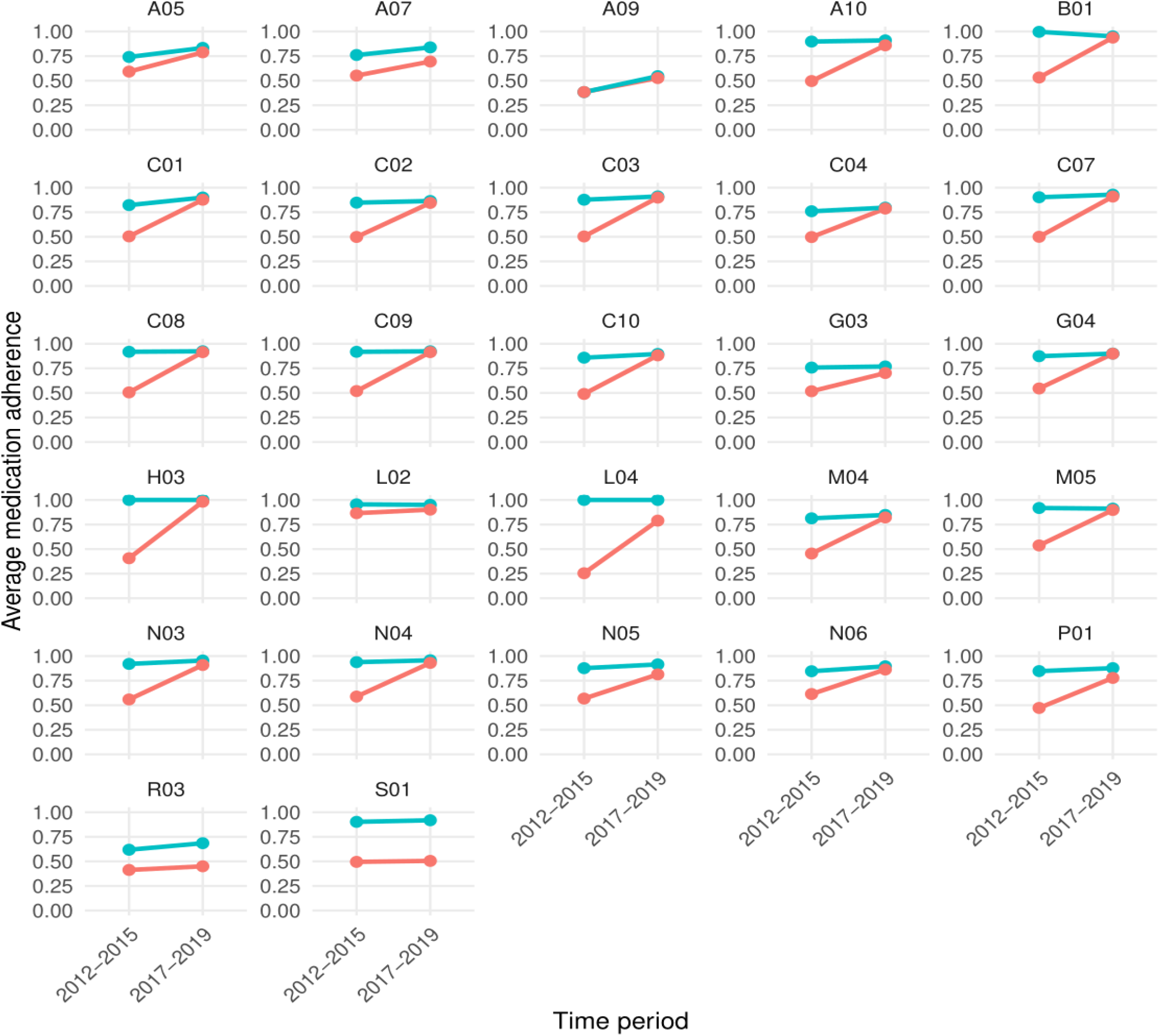
The average medication adherence by time period and ATC therapeutic subgroups. Blue – corrected dataset, red – baseline dataset. A05 - Bile and liver therapy, A07 - Antidiarrheals, intestinal antiinflammatory/antiinfective agents, A09 - Digestives, incl. enzymes, A10 - Drugs used in diabetes, B01 - Antithrombotic agents, C01 - Cardiac therapy, C02 - Antihypertensives, C03 – Diuretics, C04 - Peripheral vasodilators, C07 - Beta blocking agents, C08 - Calcium channel blockers, C09 - Agents acting on the renin-angiotensin system, C10 - Lipid modifying agents, G03 - Sex hormones and modulators of the genital system, G04 – Urologicals, H03 - Thyroid therapy, L02 - Endocrine therapy, L04 – Immunosuppressants, M04 - Antigout preparations, M05 - Drugs for treatment of bone diseases, N03 – Antiepileptics, N04 - Anti-parkinson drugs, N05 – Psycholeptics, N06 – Psychoanaleptics, P01 – Antiprotozoals, R03 - Drugs for obstructive airway diseases, S01 – Ophthalmologicals

## Discussion

### Principal Results

Accurate estimation of DS is essential for assessing medication adherence and conducting pharmacoepidemiological research. This paper set out to improve the completeness and precision of prescription data. We developed and implemented a multi-step data cleaning and imputation approach to address missing or incomplete information in prescription records, specifically targeting the derivation of DS. Unlike previous studies, which typically focused on a single drug class or drug form [4, 6-9, 11, 13-18, 20, 26, 27] our work aimed to determine DS values for all prescriptions. By applying a combination of a rule-based calculations, statistical imputation, and domain knowledge, we were able to assign DS values to almost all of the 7.5 million dispensed prescriptions included in this study dataset. This demonstrates the feasibility of using hybrid methods in large-scale, real-world prescription datasets, particularly when working with data mapped to standardized models.

Overall, the medication adherence calculated on baseline database and corrected database suggests that our approach significantly improved the adherence estimates. For most drug groups, it became evident that once imputation methods were applied, the adherence estimates for the 2012–2015 period aligned more closely with those from 2017–2019, when daily dose reporting became mandatory for physicians and the overall database quality improved. At the same time, the adherence values for 2017–2019 period for both datasets remained similar for most drug classes. This supports the hypothesis that earlier data underrepresented medication availability due to incomplete documentation, and that our imputation procedures improved temporal consistency and data reliability.

Some kind of imputation was needed in each observed drug class. The predominance of tablets, capsules and suppositories in the dataset enabled the use of daily dose based imputations for a substantial portion of the records. Imputing one dose per day has proven to be an effective way to address the missing data for some oral drugs in a previous study with hearth medications [6]. Although this approach was also applicable in our dataset, the mode tables across active substances and dosage amount level revealed that such uniform approach was unsuitable for 39.3% of single dose medications. Therefore, using mode-table imputation method helped to identify most common daily doses per active substance and use this information in DS calculations. Although our approach significantly improves the data quality, it is not flawless. For example, the medication adherence calculated for thyroid therapy (H03) on corrected database was 100%. Thus, raising the suspicion that it might be overestimated and the imputed daily doses were too small. It could be hypothesised that for some medications more than others, the individual treatment regimens may differ and therefore, it is difficult to identify the most common dose to impute. When the mode value used in imputation is lower than the next most popular value, then it can result in higher DS value and therefore better medication adherence values.

For non-single dose drug forms, such as creams, drops, and gels, imputation based on SPCs and provided DS proved useful, though these methods are inherently less precise due to variability in usage patterns and dosing recommendations. In two drug classes – obstructive airway diseases (R03) and ophthalmologicals (S01) – the medication improved for both time periods. One reason for this could be that in the baseline dataset a rough imputation of 30 days was applied to all missing daily doses while in this study the imputed DS was more active substance specific and based on national SPCs.

In addition to imputing missing data, there is sometimes a question of plausibility of provided DS that are given by doctor with the prescription [5, 19]. The question arises whether and how we assess the DS prescribed by physicians, and whether this should be compared with the calculated duration. Because until 2016 our prescription system allowed to enter the duration manually - errors can often occur. The most common and noticeable errors occurred when repeat prescriptions were issued and each was assigned a DS of 180 days, which in fact represents the combined length of three prescriptions. This stems from the common practice of doctors providing three refill prescriptions per medication. These kinds of problems were easy to notice and fix, but more complex errors were harder to detect. Another example concerns vaccines and implants, for which no standard exists for indicating the days’ supply on the prescription, and each doctor records it according to their own discretion. As a result of incorrect entries some prescriptions may end up with an inaccurate DS value. Despite these challenges, our imputation methods combined with domain knowledge ensured reasonable estimates of DS across diverse drug forms.

Our study also underscores the importance of a user design and information architecture of the prescription database. It clearly emerged from this study, that before the summer of 2016, when there was no requirement to record dosing instructions for medications, the data quality was lower and different methods were needed to backfill this information retrospectively. Therefore, to collect accurate data, more effort should be paid on the architecture of the system to ensure that all necessary data will be inserted and stored as correctly as possible. Moreover, there is a need to raise awareness of doctors on the importance of data quality and its effect on evaluating health care services and medication adherence. For example, we identified in the dataset that sometimes there is inconsistency in the units of prescribed medications within prescriptions issued by doctors – one might use units (e.g. 1 tablet) on one prescription and the amount of active substance (e.g. 10mg) on another. Moreover, some doctors have practice to renew the old prescription without changing the dosing information even when the dosing regimen changes. This all impacts the data quality as detecting such cases from the data is very difficult if not impossible. Therefore, beyond seeking sophisticated imputation methods to address the missing data, we should also consider improving the prescription systems and informing and educating the doctors, who enter this data. More complete and accurate records would provide a better foundation for secondary use of prescription data in the future.

## Limitations

One limitation of our study is, that there was no golden standard or reference database to compare the results with. Although the amount of missing data substantially reduced in period 2017-2019 due to the changes in prescription system, some inaccuracies due to human component remained. However, it could be argued that the baseline data from 2017-2019 gives considerably good indication of actual prescription patterns.

It is also important to acknowledge that imputation techniques possess inherent limitations and may not invariably produce fully accurate estimates. To construct mode table of daily doses, a certain proportion of presumably correct prescription data is required; otherwise, it cannot be compiled. Furthermore, in some drug classes where the dosing recommendations vary based on the severity of disease, the mode table did not seem to be the best approach as the medication adherence calculated on the corrected database were unrealistically high. Potentially some machine learning methods could be more effective in such case where mode tables fail, but this warrants further investigation. Moreover, machine learning methods should be also applied on other injections and less prevalent drug forms which in this study were excluded.

## Conclusion

In summary, our study demonstrated that with a carefully designed imputation pipeline where data-driven imputation is combined with domain knowledge and literature information, it is possible to meaningfully improve the quality of prescription datasets and generate more accurate and consistent adherence metrics across various drug form. Nonetheless, future efforts should continue to refine imputation techniques, incorporate machine learning approaches where appropriate, and expand validation efforts using external benchmarks or clinical outcomes.

## Data Availability

The datasets generated and analysed during this study are not publicly available due to legal restrictions on sharing de-identified data. According to legislative regulation and data protection law in Estonia, the authors cannot publicly release the data received from the health data registries in Estonia. However, the data can be requested by completing necessary applications in order to carry out research or an evaluation of public interest and acquiring the permission of the controller of the databases.

## Author Contribution

Conceptualization and methodology: MM, KM, RK, MO, JH, NU

Data curation: MM, MO, ST

Formal analysis: MM, KM

Funding and resources: RK, SR, JV Investigation: MM, KM, HK

Software: MM, JH, MO Supervision: KM, RK Visualization: MM, KM, RK

Writing – review & editing: The original draft was written by MM while all authors read and edited the manuscript and approved the final version.

### Abbreviations

ATC: Anatomical Therapeutic Chemical
DDD: defined daily dose
DS: days’ supply
CMA: medication availability measure
SPC: Summary of Product Characteristics
OMOP CDM: Observational Medical Outcomes Partnership Common Data Model

## Multimedia Appendix 1

Yearly Medication Adherence (2012–2019) showing baseline dataset and corrected dataset. Stratified by ATC code.

## References

[1] Z. Miao, M. D. Sealey, S. Sathyanarayanan, D. Delen, L. Zhu, and S. Shepherd, “A data preparation framework for cleaning electronic health records and assessing cleaning outcomes for secondary analysis,” Inf Syst, vol. 111, Jan. 2023, doi: 10.1016/j.is.2022.102130.

[2] W. M. Vollmer, M. Xu, A. Feldstein, D. Smith, A. Waterbury, and C. Rand, “Comparison of pharmacy-based measures of medication adherence,” BMC Health Serv Res, vol. 12, no. 1, 2012, doi: 10.1186/1472-6963-12-155.

[3] B. Wettermark et al., “The nordic prescription databases as a resource for pharmacoepidemiological research-a literature review,” Jul. 2013. doi: 10.1002/pds.3457.

[4] K. J. Lum et al., “Evaluation of methods to estimate missing days’ supply within pharmacy data of the Clinical Practice Research Datalink (CPRD) and The Health Improvement Network (THIN),” Eur J Clin Pharmacol, vol. 73, no. 1, pp. 115–123, Jan. 2017, doi: 10.1007/s00228-016-2148-4.

[5] C. Xu, S. A. Ferrante, T. Fitzgerald, C. D. Pericone, and B. Wu, “Inconsistencies in the days supply values reported in pharmacy claims databases for biologics with long maintenance intervals,” 2023.

[6] D. Ung et al., “Assuming one dose per day yields a similar estimate of medication adherence in patients with stroke: An exploratory analysis using linked registry data,” Br J Clin Pharmacol, vol. 87, no. 3, pp. 1089–1097, Mar. 2021, doi: 10.1111/bcp.14468.

[7] L. L. Dalli et al., “Towards better reporting of the proportion of days covered method in cardiovascular medication adherence: A scoping review and new tool TEN-SPIDERS,” Oct. 01, 2022, John Wiley and Sons Inc. doi: 10.1111/bcp.15391.

[8] P. Saukkosalmi, H. Kankaanranta, I. Vähätalo, L. Sillanmäki, and M. Sumanen, “Defined daily dose definition in medication adherence assessment in asthma,” Eur Clin Respir J, vol. 10, no. 1, 2023, doi: 10.1080/20018525.2023.2207335.

[9] S. Nielsen et al., “Defined daily doses (DDD) do not accurately reflect opioid doses used in contemporary chronic pain treatment,” Pharmacoepidemiol Drug Saf, vol. 26, no. 5, pp. 587–591, May 2017, doi: 10.1002/pds.4168.

[10] “World Health Organization. ATC/DDD Toolkit. 2020.,” https://www.who.int/tools/atc-ddd-toolkit.

[11] I. Tapply and D. C. Broadway, “Improving adherence to topical medication in patients with glaucoma,” 2021, Dove Medical Press Ltd. doi: 10.2147/PPA.S264926.

[12] S. J. Sinnott, J. M. Polinski, S. Byrne, and J. J. Gagne, “Measuring drug exposure: Concordance between defined daily dose and days’ supply depended on drug class,” J Clin Epidemiol, vol. 69, pp. 107–113, Jan. 2016, doi: 10.1016/j.jclinepi.2015.05.026.

[13] D. S. Friedman et al., “Using pharmacy claims data to study adherence to glaucoma medications: Methodology and findings of the Glaucoma Adherence and Persistency Study (GAPS),” Invest Ophthalmol Vis Sci, vol. 48, no. 11, pp. 5052–5057, Nov. 2007, doi: 10.1167/iovs.07-0290.

[14] A. S. Assem, S. A. Fekadu, A. A. Yigzaw, Z. M. Nigussie, and A. A. Achamyeleh, “Level of glaucoma drug adherence and its associated factors among adult glaucoma patients attending felege hiwot specialized hospital, bahir dar city, northwest ethiopia,” Clin Optom (Auckl), vol. 12, pp. 189–197, 2020, doi: 10.2147/OPTO.S274850.

[15] P. Muñoz-Villegas, H. Martínez-Bautista, and O. Olvera-Montaño, “Determinants of adherence to treatment in patients with ophthalmic conditions,” Expert Rev Clin Pharmacol, vol. 16, no. 12, pp. 1249–1259, 2023, doi: 10.1080/17512433.2023.2279740.

[16] G. Gupta, P. Mallefet, D. W. Kress, and A. Sergeant, “Adherence to topical dermatological therapy: Lessons from oral drug treatment,” 2009. doi: 10.1111/j.1365-2133.2009.09253.x.

[17] C. Fenske, N. Boytsov, J. Guo, and Z. Dawson, “Prescription Market Share and Treatment Patterns in Atopic Dermatitis: A Retrospective Observational Study Using US Insurance Claims,” Adv Ther, vol. 39, no. 5, pp. 2052–2064, May 2022, doi: 10.1007/s12325-022-02071-y.

[18] A. Lo, K. K. Lovell, J. D. Greenzaid, M. E. Oscherwitz, and S. R. Feldman, “Adherence to treatment in dermatology: Literature review,” JEADV Clinical Practice, vol. 3, no. 2, pp. 401–418, Jun. 2024, doi: 10.1002/jvc2.379.

[19] H. Singh, S. Mani, D. Espadas, N. Petersen, V. Franklin, and L. A. Petersen, “Prescription Errors and Outcomes Related to Inconsistent Information Transmitted Through Computerized Order Entry A Prospective Study.”

[20] A. M. Burden, A. Huang, M. Tadrous, and S. M. Cadarette, “Variation in the days supply field for osteoporosis medications in Ontario,” Arch Osteoporos, vol. 8, no. 1–2, Dec. 2013, doi: 10.1007/s11657-013-0128-1.

[21] M. Oja et al., “Transforming Estonian health data to the Observational Medical Outcomes Partnership (OMOP) Common Data Model: Lessons learned,” JAMIA Open, vol. 6, no. 4, Dec. 2023, doi: 10.1093/jamiaopen/ooad100.

[22] “The Register of Medicinal Products,” https://www.ravimiregister.ee/.

[23] “Medicinal Products Act,” https://www.riigiteataja.ee/akt/130042025002?leiaKehtiv.

[24] A. L. Dima and D. Dediu, “Computation of adherence to medication and visualization of medication histories in R with AdhereR: Towards transparent and reproducible use of electronic healthcare data,” PLoS One, vol. 12, no. 4, Apr. 2017, doi: 10.1371/journal.pone.0174426.

[25] J. Holm, “AdherenceFromOmop,” https://gitlab.cs.ut.ee/johanneh/AdherenceFromOMOP.

[26] A. J. Nelson, N. J. Pagidipati, and H. B. Bosworth, “Improving medication adherence in cardiovascular disease,” Jun. 01, 2024, Nature Research. doi: 10.1038/s41569-023-00972-1.

[27] H. Tibble, A. Sheikh, and A. Tsanas, “Estimating medication adherence from Electronic Health Records: comparing methods for mining and processing asthma treatment prescriptions,” BMC Med Res Methodol, vol. 23, no. 1, Dec. 2023, doi: 10.1186/s12874-023-01935-3.

