## Appendix 1 for "A Comprehensive Approach to Days’ Supply Estimation in a Real-World Prescription Database: Data Cleaning, Imputation, and Adherence Analysis"

**Figure S1:** Graphs of yearly medication adherence (2012–2019) showing baseline dataset and corrected dataset. Stratified by ATC code.

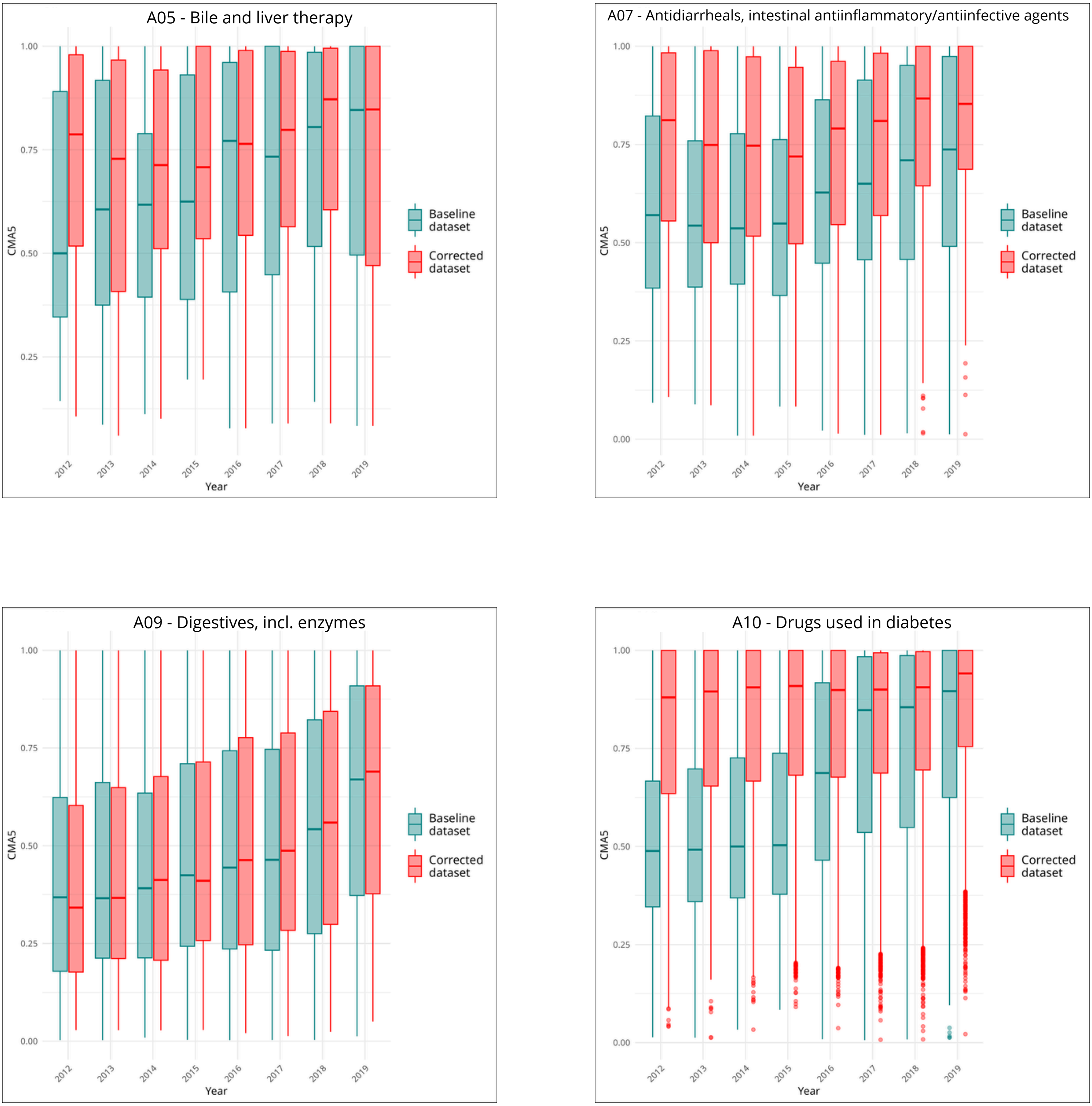

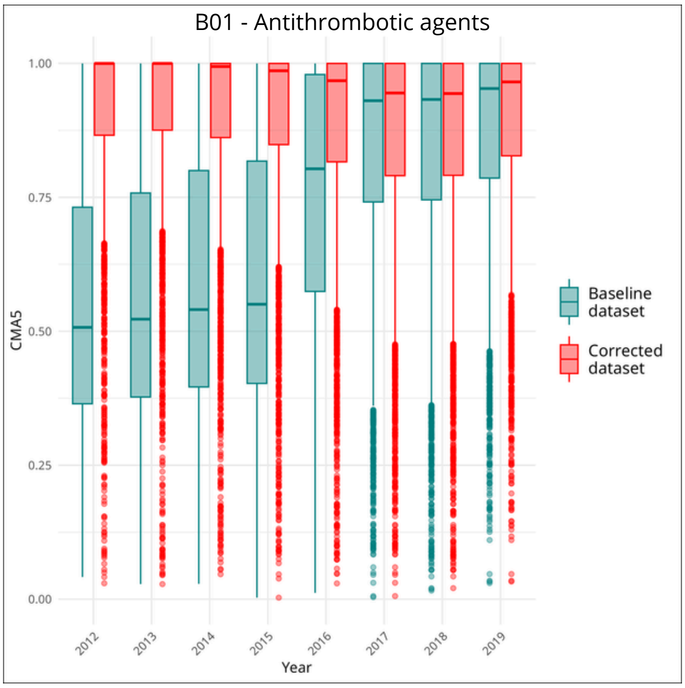
B01 - Antithrombotic agents
CMA5
1.00
0.75
0.50
0.25
0.00
2012
2013
2014
2015
2016
2017
2018
2019
Year
Baseline dataset
Corrected dataset

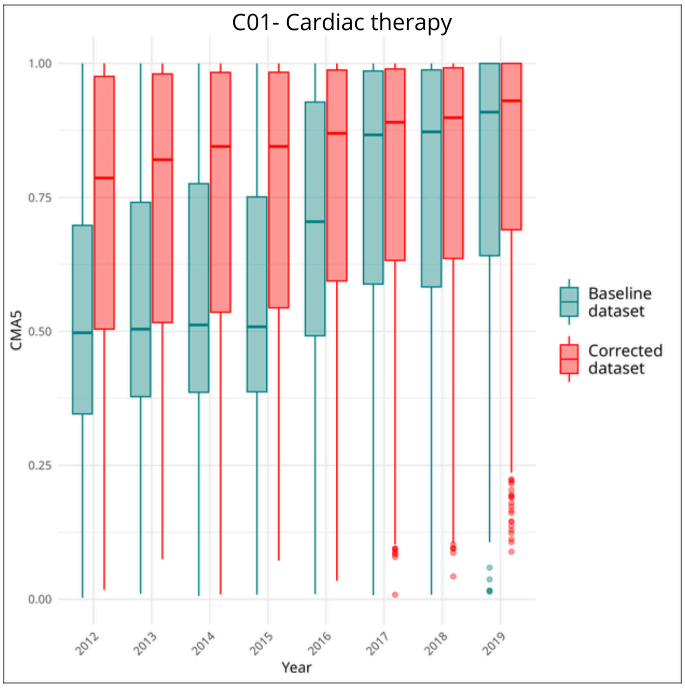
C01- Cardiac therapy
CMA5
1.00
0.75
0.50
0.25
0.00
2012
2013
2014
2015
2016
2017
2018
2019
Year
Baseline dataset
Corrected dataset

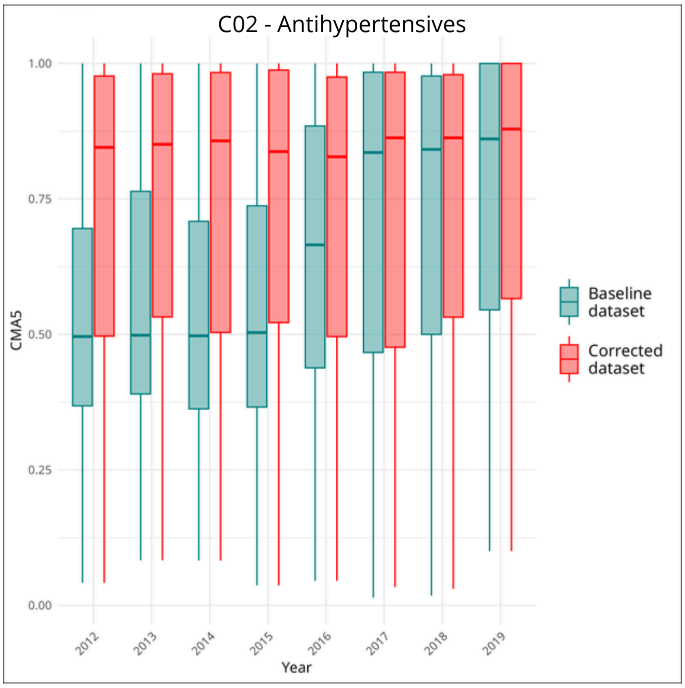
C02 - Antihypertensives
CMA5
1.00
0.75
0.50
0.25
0.00
2012
2013
2014
2015
2016
2017
2018
2019
Year
Baseline dataset
Corrected dataset

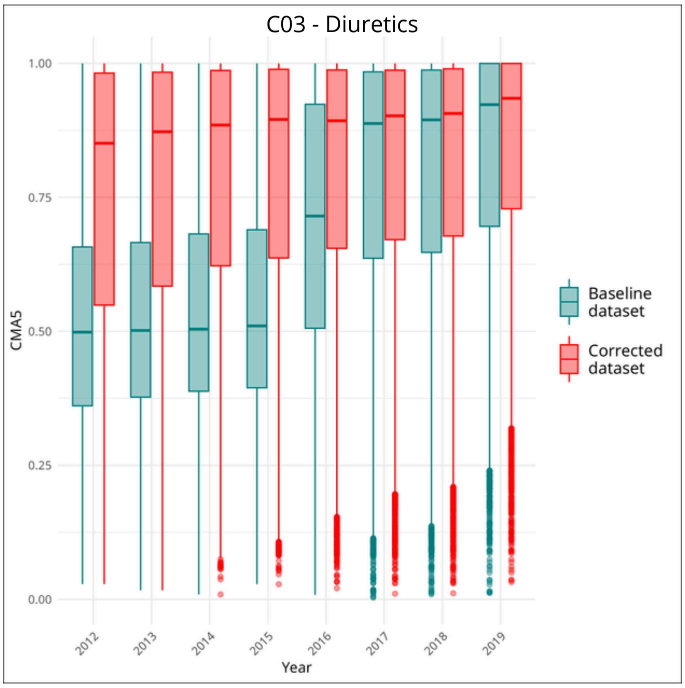
C03 - Diuretics
CMA5
1.00
0.75
0.50
0.25
0.00
2012
2013
2014
2015
2016
2017
2018
2019
Year
Baseline dataset
Corrected dataset

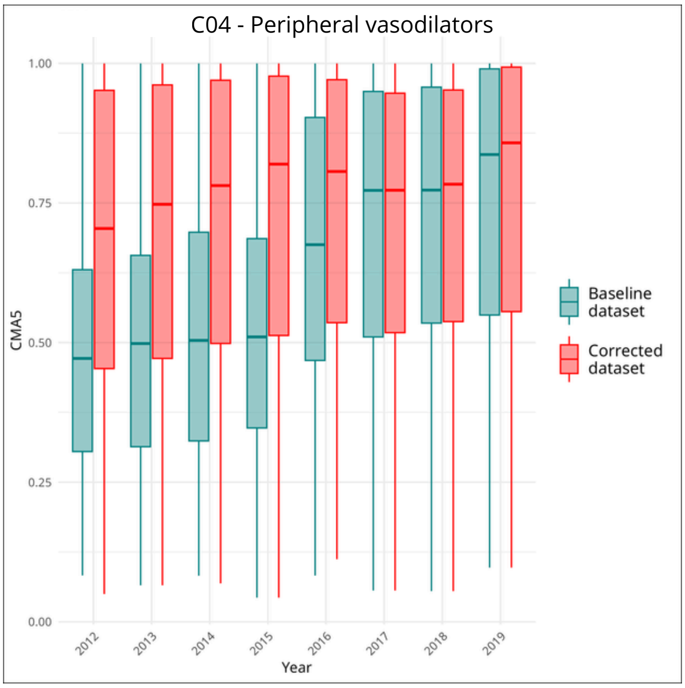
C04 - Peripheral vasodilators
CMA5
1.00
0.75
0.50
0.25
0.00
2012
2013
2014
2015
2016
2017
2018
2019
Year
Baseline dataset
Corrected dataset

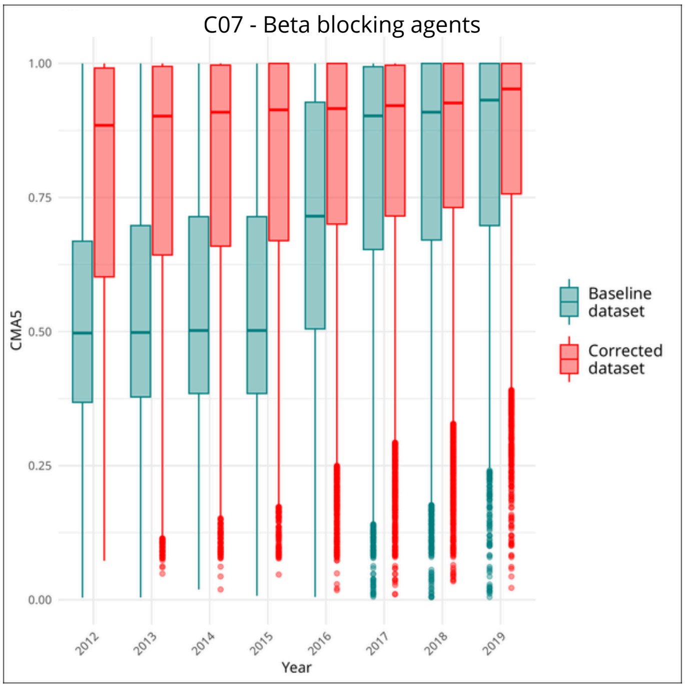
C07 - Beta blocking agents
CMA5
1.00
0.75
0.50
0.25
0.00
2012
2013
2014
2015
2016
2017
2018
2019
Year
Baseline dataset
Corrected dataset

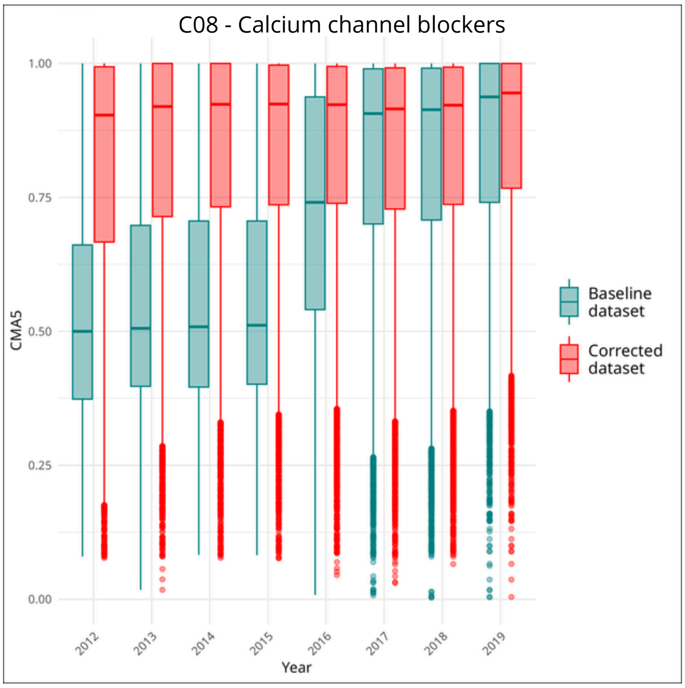
C08 - Calcium channel blockers
CMA5
Year
Baseline dataset
Corrected dataset

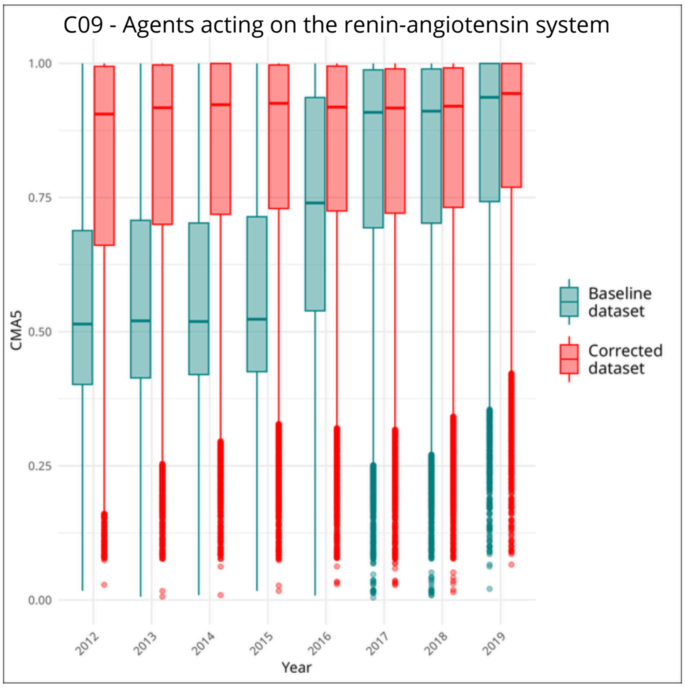
C09 - Agents acting on the renin-angiotensin system
CMA5
Year
Baseline dataset
Corrected dataset

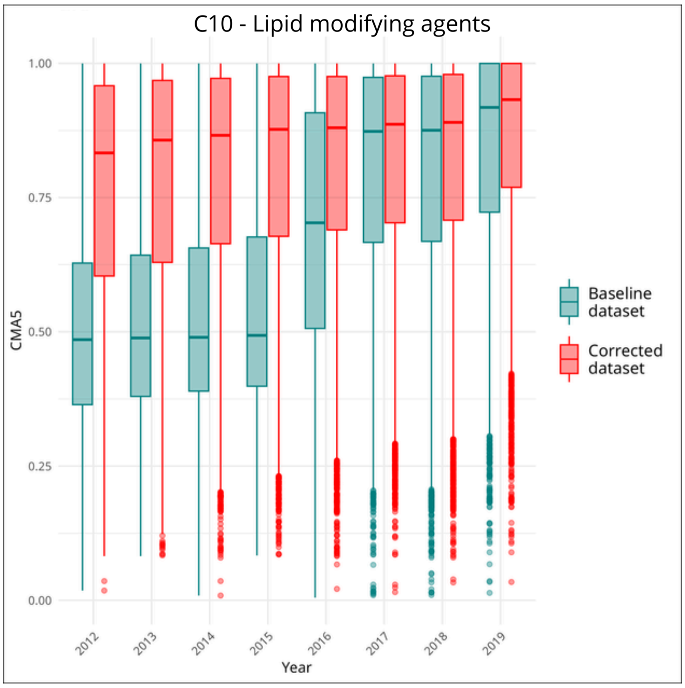
C10 - Lipid modifying agents
CMA5
Year
Baseline dataset
Corrected dataset

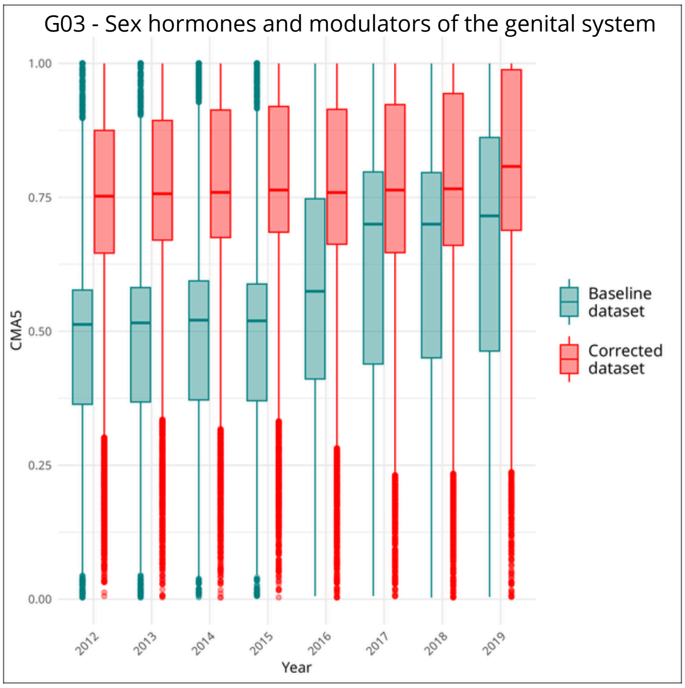
G03 - Sex hormones and modulators of the genital system
CMA5
Year
Baseline dataset
Corrected dataset

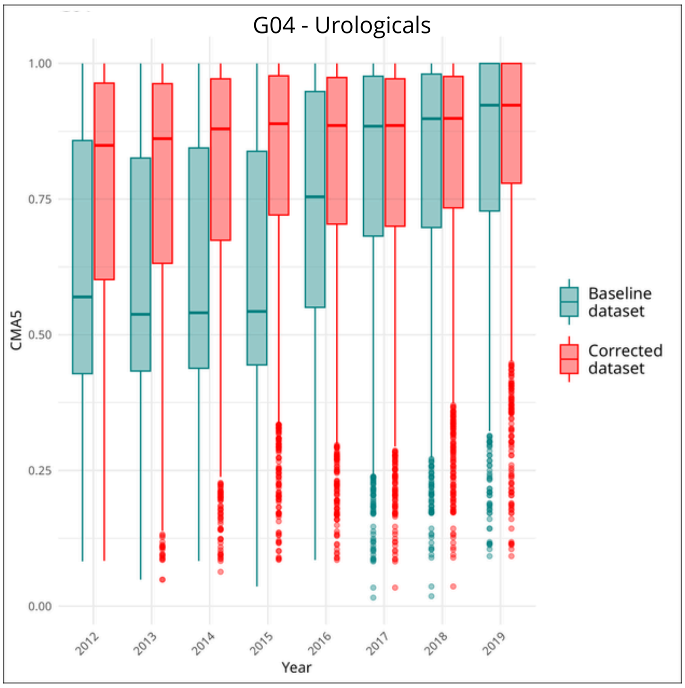
G04 - Urologicals
CMA5
Year
Baseline dataset
Corrected dataset

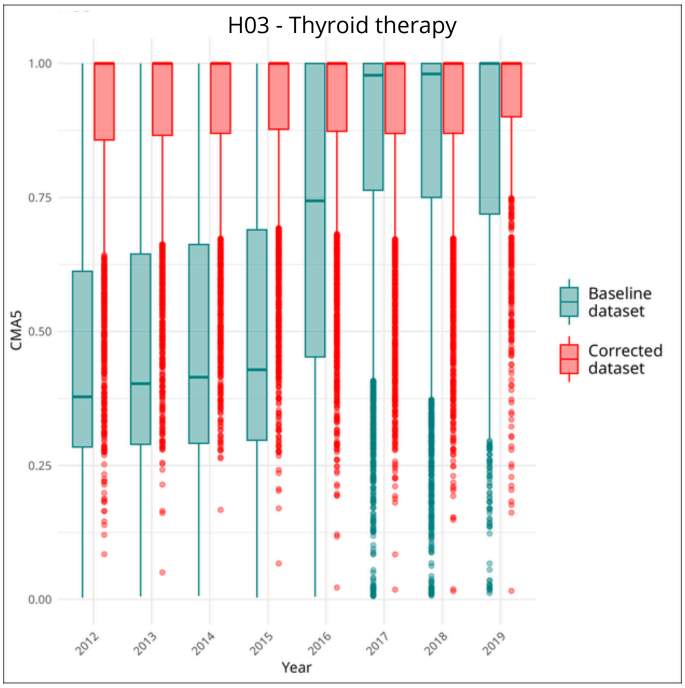
H03 - Thyroid therapy
CMA5
Year
Baseline dataset
Corrected dataset

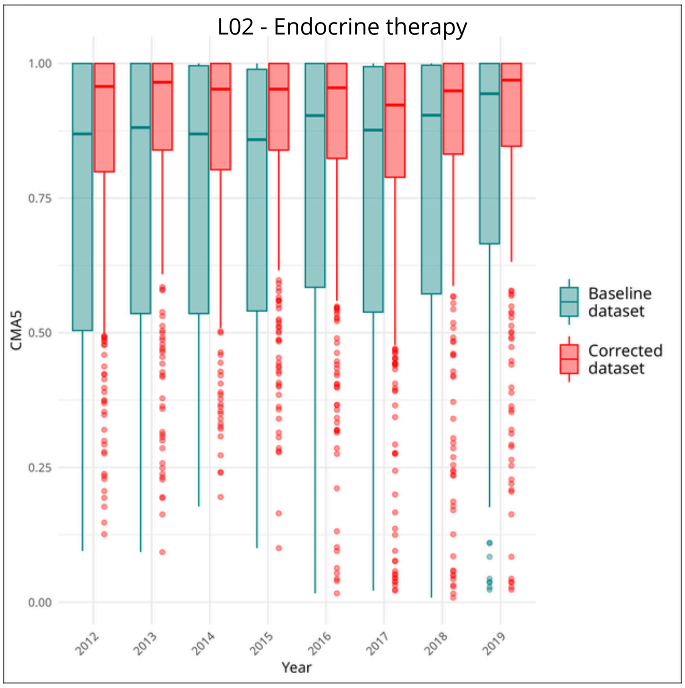
L02 - Endocrine therapy
1.00
0.75
0.50
0.25
0.00
CMA5
2012
2013
2014
2015
2016
2017
2018
2019
Year
Baseline dataset
Corrected dataset

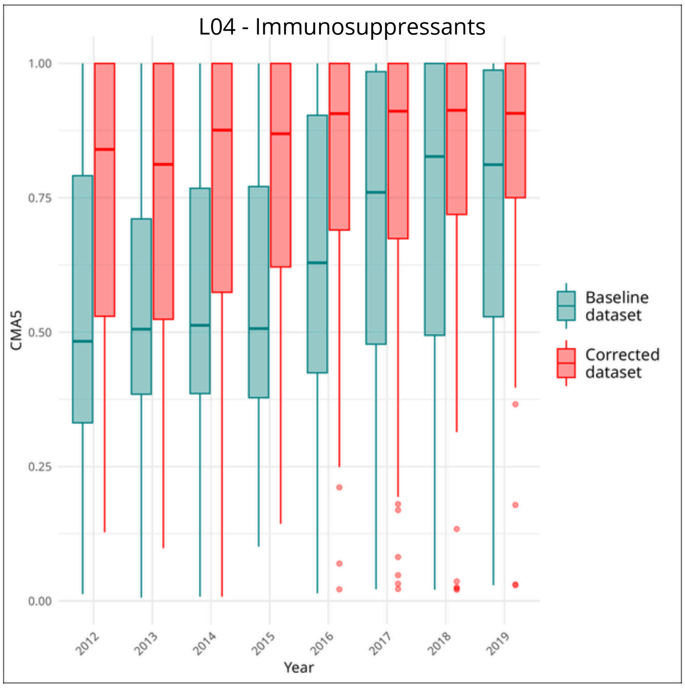
L04 - Immunosuppressants
1.00
0.75
0.50
0.25
0.00
CMA5
2012
2013
2014
2015
2016
2017
2018
2019
Year
Baseline dataset
Corrected dataset

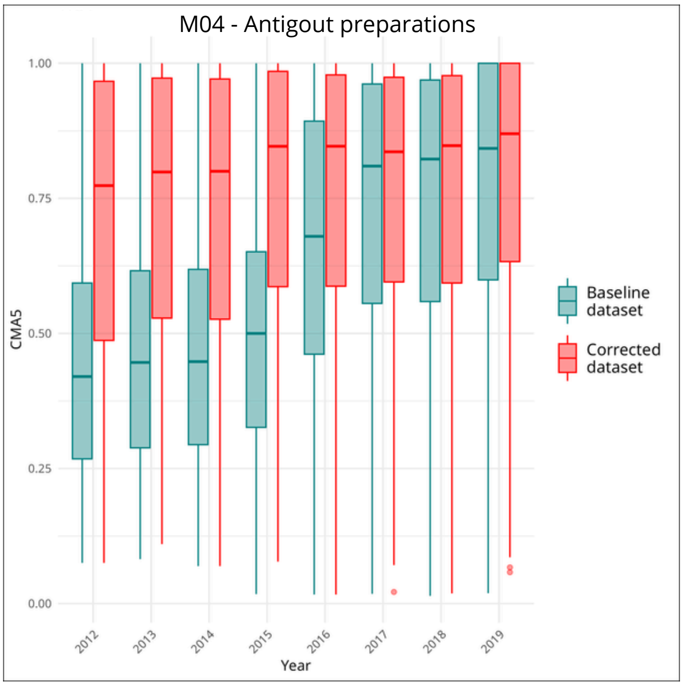
M04 - Antigout preparations
1.00
0.75
0.50
0.25
0.00
CMA5
2012
2013
2014
2015
2016
2017
2018
2019
Year
Baseline dataset
Corrected dataset

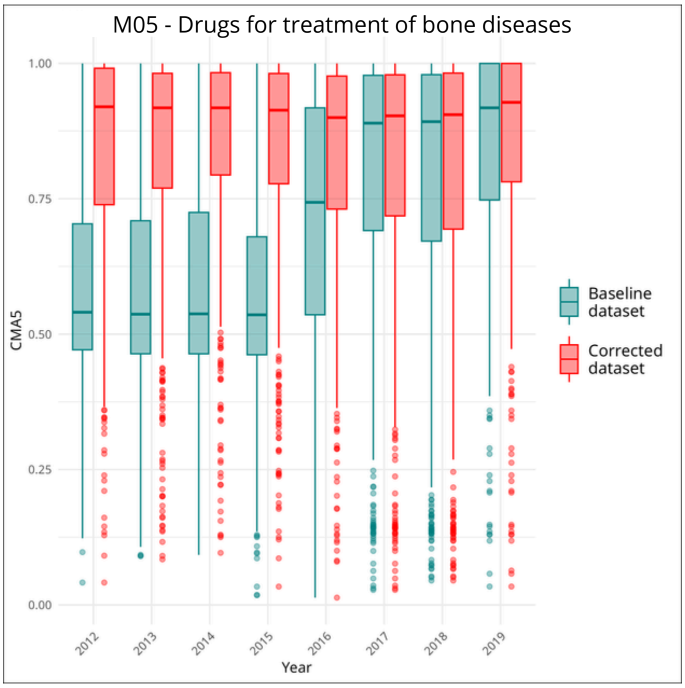
M05 - Drugs for treatment of bone diseases
1.00
0.75
0.50
0.25
0.00
CMA5
2012
2013
2014
2015
2016
2017
2018
2019
Year
Baseline dataset
Corrected dataset

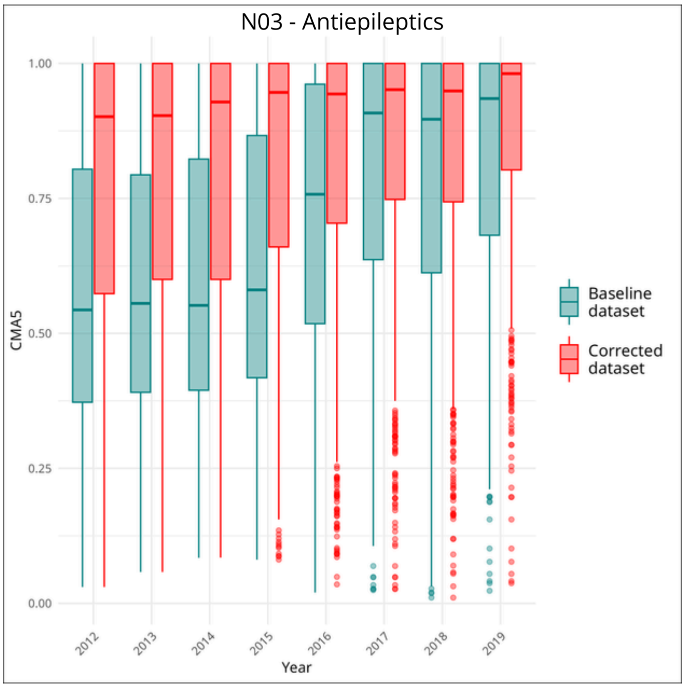
N03 - Antiepileptics
1.00
0.75
0.50
0.25
0.00
CMA5
2012
2013
2014
2015
2016
2017
2018
2019
Year
Baseline dataset
Corrected dataset

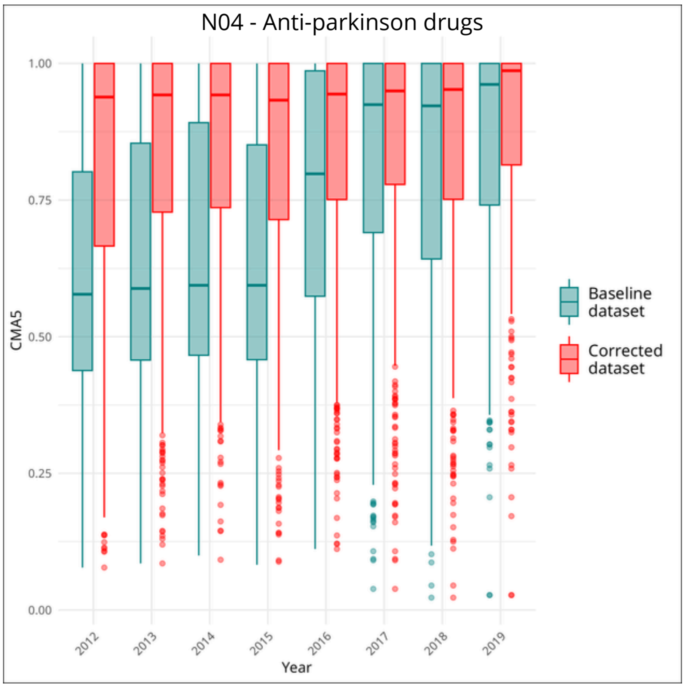
N04 - Anti-parkinson drugs
1.00
0.75
0.50
0.25
0.00
CMA5
2012
2013
2014
2015
2016
2017
2018
2019
Year
Baseline dataset
Corrected dataset

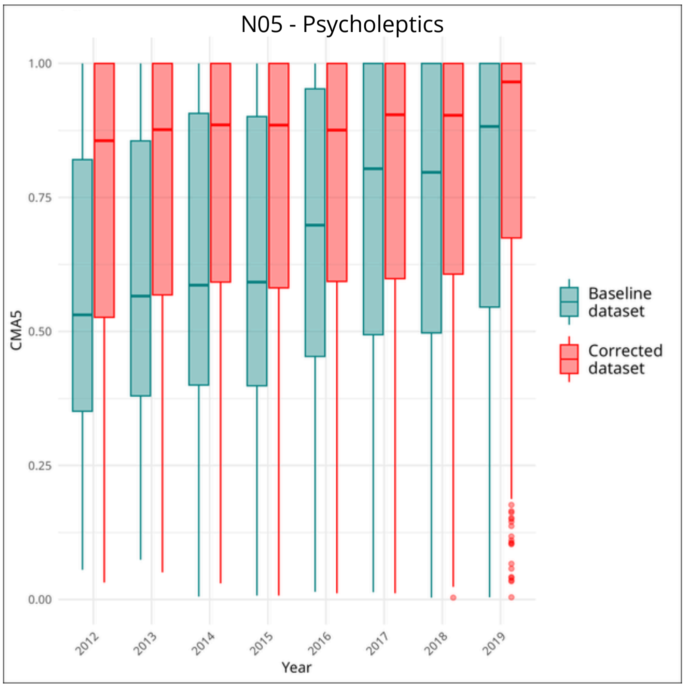
N05 - Psycholeptics
CMA5
1.00
0.75
0.50
0.25
0.00
2012
2013
2014
2015
2016
2017
2018
2019
Year
Baseline dataset
Corrected dataset

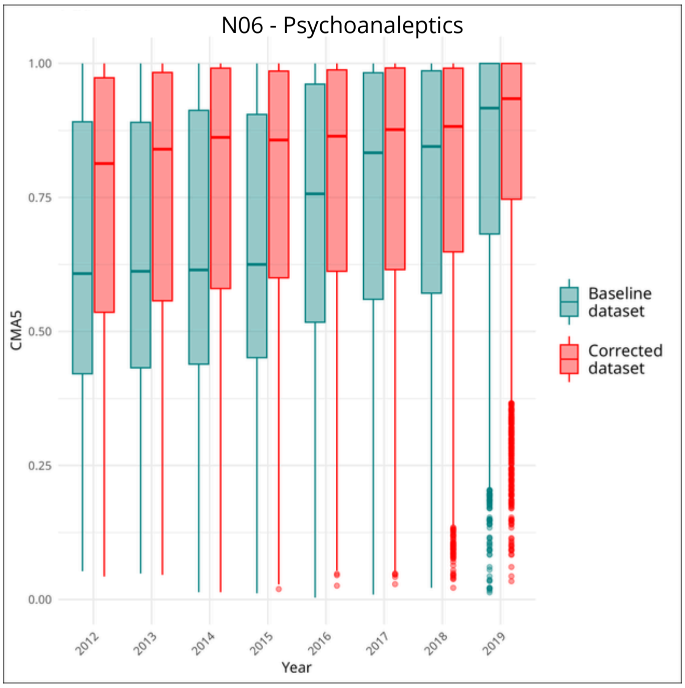
N06 - Psychoanaleptics
CMA5
1.00
0.75
0.50
0.25
0.00
2012
2013
2014
2015
2016
2017
2018
2019
Year
Baseline dataset
Corrected dataset

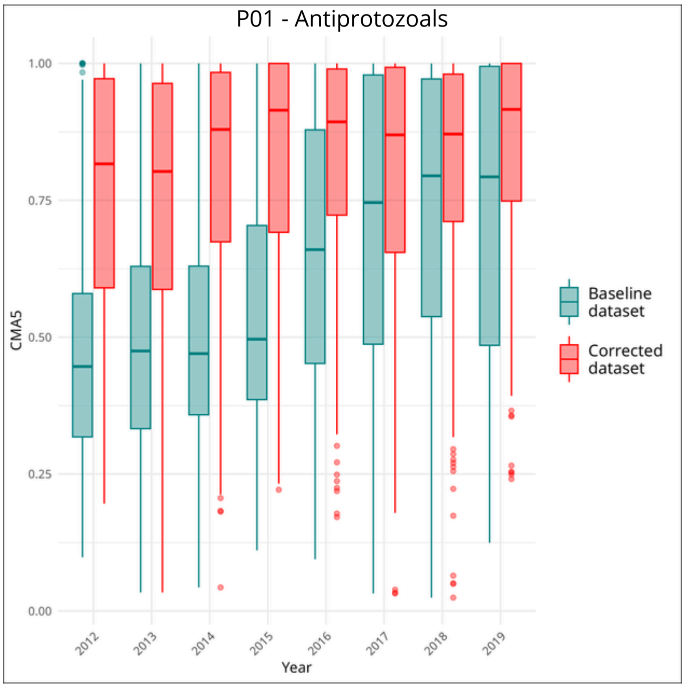
P01 - Antiprotozoals
CMA5
1.00
0.75
0.50
0.25
0.00
2012
2013
2014
2015
2016
2017
2018
2019
Year
Baseline dataset
Corrected dataset

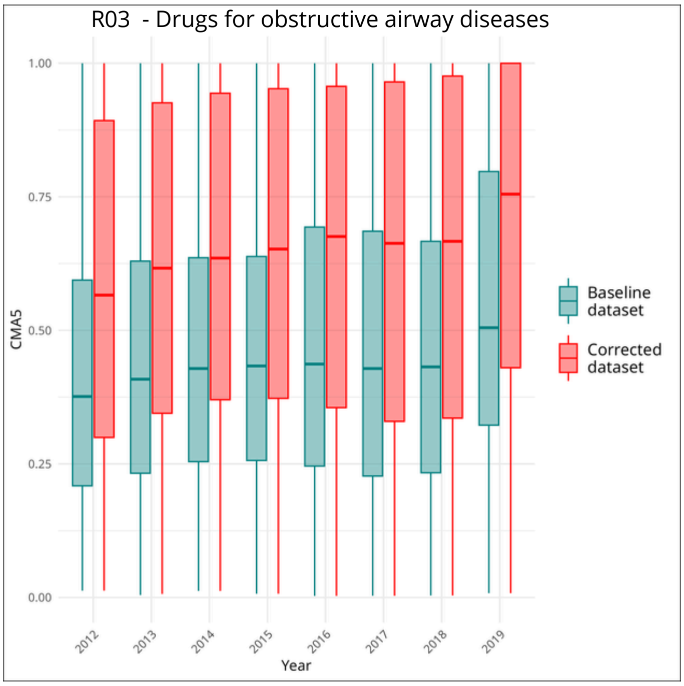
R03 - Drugs for obstructive airway diseases
CMA5
1.00
0.75
0.50
0.25
0.00
2012
2013
2014
2015
2016
2017
2018
2019
Year
Baseline dataset
Corrected dataset

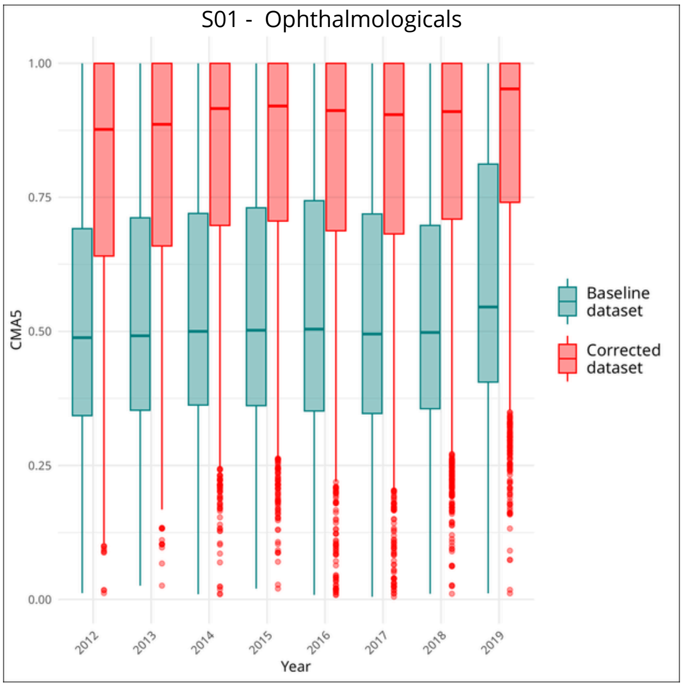
S01 - Ophthalmologicals
CMA5
1.00
0.75
0.50
0.25
0.00
2012
2013
2014
2015
2016
2017
2018
2019
Year
Baseline dataset
Corrected dataset
